# Characteristics of lymphocyte subsets and cytokines in peripheral blood of 123 hospitalized patients with 2019 novel coronavirus pneumonia (NCP)

**DOI:** 10.1101/2020.02.10.20021832

**Authors:** Suxin Wan, Qingjie Yi, Shibing Fan, Jinglong Lv, Xianxiang Zhang, Lian Guo, Chunhui Lang, Qing Xiao, Kaihu Xiao, Zhengjun Yi, Mao Qiang, Jianglin Xiang, Bangshuo Zhang, Yongping Chen

## Abstract

**Background:** To explore the cellular immunity and cytokines status of NCP patients and to predict the correlation between the cellular immunity levels, cytokines and the severity of patients.

**Methods:** 123 NCP patients were divided into mild and severe groups. Peripheral blood was collected, lymphocyte subsets and cytokines were detected. Correlation analysis was performed on the lymphocyte subsets and cytokines, and the differences between the indexes of the two groups were analyzed.

**Results:** 102 mild and 21 severe patients were included. Lymphocyte subsets were reduced in two groups. The proportion of CD8 + T reduction in the mild and severe group was 28.43% and 61.9%, respectively; The proportion of B cell reduction was 25.49% and 28.57%; The proportion of NK cell reduction was 34.31% and 47.62%; The detection value of IL-6 was 0 in 55.88% of the mild group, mild group has a significantly lower proportion of patients with IL-6 higher than normal than severe group; There was no significant linear correlation between the lymphocyte subsets and cytokines, while significant differences were noticed between the two groups in CD4 + T, CD8 + T, IL-6 and IL-10.

**Conclusions:** Low levels of CD4+T and CD8+T are common in severe NCP. IL-6 and IL-10 levels were higher in severe patients. T cell subsets and cytokines can be used as one of the basis for predicting the transition from mild to severe. Large number of samples are still needed to confirm the “warning value” of CD4 + T, CD8 + T IL-6 and IL-10.

## Introduction

2019-novel coronavirus (2019-nCoV), which was discovered due to viral pneumonia cases in Wuhan in 2019, was named by the World Health Organization (WHO) on 10 January 2020. The main source of infection is pneumonia patients infected by new coronavirus. Respiratory droplets are the main route of transmission and it can also be transmitted through contact. People are generally susceptible. At present, the occurrence, development, mechanism of prognosis and immune status of patients with 2019-nCoV are still unclear.^1^ This study intends to detect the peripheral blood lymphocyte subsets and cytokines in 123 patients with 2019-nCoV infection by using four-color immunoflow cytometry and multiple microsphere flow immunofluorescence to investigate the cellular immunity status and cytokine levels of 2019-nCoV infected patients, to explore the correlation between cellular immune statuses, cytokine levels and the severity of 2019-nCoV infection.

## Materials and Methods

### Patients

From January 26, 2019 to February 4, 2020, 123 inpatients diagnosed with 2019-nCoV infection were collected in the Chongqing Three Gorges Central Hospital. All patients were confirmed to be positive for new coronavirus nucleic acid by real-time fluorescent RT-PCR. Patients were diagnosed according to the World Health Organization interim guidance for NCP and divided into mild and severe (including severe and critical) groups

### Blood sampling

Blood samples of the patients were collected by the nurse according to the doctor’s order, and all patients were not treated before the blood sampling or did not receive the standardized treatment according to the diagnosis and treatment scheme of NCP.

### Lymphocyte subsets detection

Two test tubes were taken for each patient, and were numbered A and B respectively. 5 μL of CD3/ CD8/ CD45/ CD4 antibody (Beijing Tongshengshidai Biotechnology Co., Ltd) was added to the tube A and 5 μL of CD16 + 56/ CD45/ CDl9 antibody was added to tube B. After adding 50 μL of EDTA anticoagulant whole blood to each tube, the tubes were vortexed and kept at room temperature for 15 minutes in the dark. Then the samples were detected by four-color fluorescence labeled flow cytometry (Mindray BriCyte E6; Mindray, Shenzhen, China).

### Cytokine detection

The detection reagent of cytokine was provided by Qingdao Raisecare Biotechnology Co., Ltd (calibrator lot number: 20190801). Six cytokines including IL-4, IL-6, IL-10, IL-17, TNF and IFN were detected by multiple microsphere flow immunofluorescence according to the manufacturer’s instructions. After the blood sample and the corresponding flow tube be numbered 101, 102, 103, 104, and 105, EDTA-K2 anticoagulant whole blood was centrifuged at 2365 r / min for 30 min. Then 25 μL of experimental buffer, 25 μL of centrifuged plasma, 25 μL of capture microsphere antibody, and 25 μL of detection antibody were added to the corresponding flow tube. After incubating at room temperature for 2 hours in the dark with gentle shaking, 25 μL SA-PE was added into the flow tube respectively, and then incubation was continued for 30 minutes. Subsequently, the diluted wash buffer (1:10) was added to the flow tube. After a few seconds of vortex shaking, the flow tube was centrifuged at 1500r / min for 5 minutes, the liquid was slowly poured out, and the flow tube was inverted on the absorbent paper. Then 100 μL of diluted washing buffer (1:10) was added to the flow tube according to the requirements of the flow cytometer, and the test was performed after shaking for 10 seconds.

### Statistical analysis

All statistical analyses were performed using SPSS 22.0 (SPSS Inc., Chicago, IL, USA). Descriptive analyses were performed for categorical variables such as gender. Continuous variables such as inspection results were expressed as 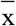 ± s and compared using the independent samples t-test. Correlation analysis results were expressed by Pearson correlation coefficient and a larger r^2^ indicates better linear correlation. P<0.05 was considered as statistically significant.

### Ethics approval

This study was approved by the ethical committee of Chongqing Three Gorges Central Hospital and informed consent was obtained from each patient.

## Results

### Baseline data

The study population included 138 hospitalized patients with confirmed NCP. 102 NCP patients (55 males and 47 females) with a mean age of 43.05 ± 13.12 (15∼82) years in the mild group and 21 NCP patients (11 males and 10 females) with a mean age of 61.29±15.55 (34∼79) years in the severe group were enrolled.

### Lymphocyte subsets in peripheral blood of NCP patients

According to the results of each index, CD4 + T, CD8 + T, B cell, NK cell, CD4 + T / CD8 + T were divided into below normal value, within normal value and above normal value. The corresponding quantities and proportions were respectively calculated, and the results were shown in **Table 1**.

**Table 1.** Distribution of lymphocyte subsets in peripheral blood of 123 patients with novel coronavirus (2019-nCoV) pneumonia (NCP)

| groups | groups | CD4<sup>+</sup>T | CD8<sup>+</sup>T | B Cell | NK Cell | CD4<sup>+</sup>T / CD8<sup>+</sup>T |
| --- | --- | --- | --- | --- | --- | --- |
| Mild group | below normal values (n, %) | 54(52.90) | 29(28.40) | 26(25.49) | 35(34.31) | 3(2.94) |
|  | within normal values (n, %) | 48(47.10) | 73(71.60) | 75(73.50) | 66(64.71) | 95(93.14) |
|  | above normal values (n, %) | - | - | 1(0.01) | 1(0.01) | 4(3.92) |
| severe group | below normal values (n, %) | 20(95.24) | 13(61.90) | 6(28.57) | 10(47.62) | 2(9.53) |
|  | within normal values (n, %) | 1(4.76) | 8(38.10) | 15(71.43) | 11(52.38) | 18(85.71) |
|  | above normal values (n, %) | - | - | - | - | 1(4.76) |

Among the mild patients, 54 (52.90%) patients had CD4 + T below normal values, 48 (47.10%) were within normal values; 73 (71.60%) patients had CD8 + T within normal values, and 29 (28.40%) were lower than normal; 26 (25.49%) patients had B cell lower than normal, 75 (73.50%) were within normal, 1 (0.01%) was higher than normal; 35 (34.31%) patients had NK cell below normal values, 66 cases (64.71%) were within normal values, 1 case (0.01%) was higher than normal values; 95 patients (93.14%) had CD4 + T / CD8 + T within normal values, 3 cases (2.94%) were lower than normal, 4 cases (3.92%) were higher than normal values.

In severe patients, CD4 + T was lower than normal in 20 patients (95.24%), within normal in 1 patient (4.76%); CD8 + T was lower than normal in 13 patients (61.90%), within normal in 8 patients (38.10%); B cell was lower than normal in 6 patients (28.57%), within normal in 15 patients (71.43%); NK cell was lower than normal in 10 patients (47.62%), and within normal in 11 patients (52.10%) 38%, 18 (85.71%) patients had CD4 + / CD8 + ratio within the normal value, 2 (9.53%) patients were lower than the normal value, 1 (4.76%) was higher than the normal value.

### Cytokines in peripheral blood of NCP patients

IL-4, IL-6, IL-10, IL-17, TNF, and IFN were divided into values of 0, within the normal value (normal values other than 0), and above the normal value according to the results of various indicators. The corresponding quantities and proportions were counted (**Table 2**).

**Table 2.** Cytokines status in peripheral blood of 123 patients with novel coronavirus (2019-nCoV) pneumonia (NCP)

| groups | groups | IL-4 | IL-6 | IL-10 | IL-17 | TNF | IFN |
| --- | --- | --- | --- | --- | --- | --- | --- |
| Mild group | 0 (n, %) | - | 57(55.88) | - | - | - | 5(4.90) |
|  | within normal values (n, %) | 102(100.00) | 14(13.73) | 102(100.00) | 102(100.00) | 100(98.04) | 92(90.20) |
|  | above normal values (n, %) | - | 31(30.39) | - | - | 2(1.96) | 5(4.90) |
| severe group | 0 (n, %) | - | 3(14.29) | - | - | - | - |
|  | within normal values (n, %) | 21(100.00) | 2(9.52) | 21(100.00) | 21(100.00) | 21(100.00) | 20(95.24) |
|  | above normal values (n, %) | - | 16(76.19) | - | - | - | 1(4.76) |

Among the mild patients, 102 (100%) patients had IL-4, IL-10, and IL-17 all within normal values; 57 (55.88%) patients had IL-6 values of 0 and 14 (13.73%) within normal values, 31 (30.39%) were higher than normal; 100 (98.04%) patients had TFN values within normal values, 2 (1.96%) were higher than normal values; 92 (90.20%) patients had IFN within the normal values, 5 cases (4.90%) had IFN values of 0, and 5 cases (4.90%) were higher than normal values.

In severe patients, IL-4, IL-10, IL-17 and TNF were all within the normal values in 21 (100%) patients; IL-6 was 0 in 3 patients (14.29%), 2 patients (9.52%) were within the normal values, 16 patients (76.19%) were higher than the normal values; IFN was within the normal values in 20 patients (95.24%), and 1 patient (4.76%) was higher than the normal values.

### Correlation between lymphocyte subsets and IL-6 in NCP patients

Considering that the value of IL-6 changed most in the mild and severe groups, the correlation between the lymphocyte subsets and the cytokine IL-6 was analyzed. The data with IL-6 value of 0 in each group (57 cases in light group and 3 cases in severe group) were excluded, and 45 cases in the mild group and 18 cases in severe group were included in the correlation analysis. As shown in **Table 3**, the correlation analysis between IL-6 and lymphocyte subsets showed that the Pearson correlation coefficients of IL-6 and CD4 + T, CD8 + T, B cell, NK cell, CD4 + T / CD8 + T were very low, and there was no significant linear correlation.

**Table 3.** Correlation analysis between IL-6 and peripheral blood lymphocytes

|  |  |  |  |  |  |
| --- | --- | --- | --- | --- | --- |
| the Mild group (n=45) |  |  |  |  |  |
| lymphocyte subsets | CD4<sup>+</sup>T | CD8<sup>+</sup>T | B Cell | NK Cell | CD4<sup>+</sup>T / CD8<sup>+</sup>T |
| r<sup>2</sup> | 0.00000325 | 0.00098 | 0.0025 | 0.0022 | 0.0003 |
| the severe group (n=18) |  |  |  |  |  |
| lymphocyte subsets | CD4<sup>+</sup>T | CD8<sup>+</sup>T | B Cell | NK Cell | CD4<sup>+</sup>T / CD8<sup>+</sup>T |
| r<sup>2</sup> | 0.04561 | 0.0445 | 0.05237 | 0.0005074 | 0.05673 |

### Comparison of lymphocyte subsets and cytokines in peripheral blood between patients with mild and severe NCP

The data (57 cases of IL-6 and 3 cases of IFN in the mild group) with the index value of 0 in the two groups were excluded. Two independent-samples t test was performed on the lymphocyte subsets and cytokines of the mild group and the severe group, with α= 0.05 as the inspection level. Significant differences were observed in CD4 + T, CD8 + T, IL-6, IL-10 between the mild group and the severe group (P < 0.05), while no significant difference was detected in B cell, NK cell, CD4 + T / CD8 + T, IL-4, IL-17, TNF, IFN between the two groups (P > 0.05, **Table 4**).

**Table 4.**
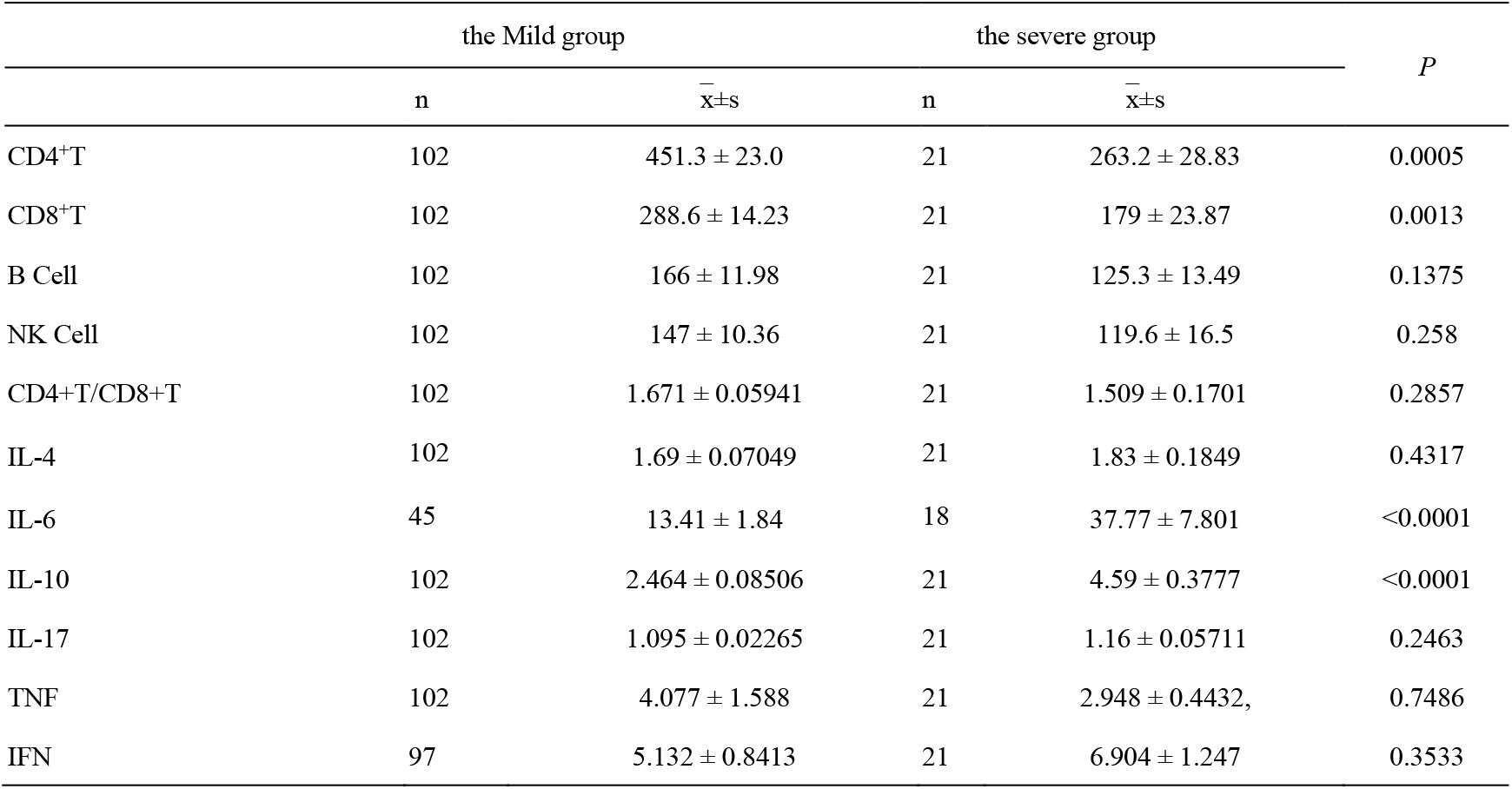
Comparison of lymphocyte subsets and cytokines in peripheral blood between mild and severe patients

## Discussion

Lymphocyte subsets play an important role in the body’s cellular immune regulation, and each cell restricts and regulates each other. This study found that among NCP patients, the reduction rate of CD4 + T accounted for 52.90% in the mild group, and 95.24% in the severe group; the reduction rate of CD8 + T accounted for 28.40% in the mild group, and 61.90% in the severe group, indicating that T lymphocytes were more inhibited in severe patients when the body is resistant to 2019-nCoV infection. This was consistent with the research conclusions of Huabiao Chen^2^ in SARS coronavirus, revealing that the body responds in the same way when coping with homologous coronavirus infection. The reduction ratio of B cell was 25.49% and 28.57% in the mild group and the severe group, respectively, with no significant difference. The reduction ratio of NK cells accounted for 34.31% in the mild group and 47.62% in the severe group, which suggested that 2019-nCoV infection limited the activity of NK cells to a certain extent, and in view of the fact that immune adjuvant IL-2 can improve the activity of NK cells, the above research results may provide new ideas and evidence for clinical treatment. ^3^

Chaolin Huang et al.^4^ showed that the levels of IL-2, IL-7, IL-10, TNF-α, G-CSF, IP-10, MCP-1, MIP-1A were significantly higher in 2019-nCoV infected patients in ICU than those in non-ICU patients, and the incidence of ARDS, secondary infection, shock, acute heart, and kidney injury was significantly higher in patients with ICU than in non-ICU patients. The results of this study indicated that 21 patients in the severe group had IL-4, IL-10, IL-17, and TNF within normal values. TNF and IFN mainly secreted by Th1 cells and NK cells were not significantly different between the mild and severe groups. In the mild group, 55.88% of the patients had IL-6 detection value of 0, and 30.39% of the patients had IL-6 detection value higher than the normal value. The reason remains to be elucidated, which may be related to the inhibition of Th2 cells involved in humoral immunity in the early stage of infection. However, the proportion of IL-6 above normal was 76.19% in the severe group, which was significantly higher than that in the mild group. This is in line with the concept of “Cytokine Storm”, which must be experienced by patients with mild illness to become severe, emphasized by Lanjuan Li, an academician of the Chinese Academy of Engineering. In addition, in the course of diagnosis and treatment, the Chongqing Three Gorges Central Hospital noticed that monitoring the changes of cytokines during the treatment process is of certain significance to optimize the treatment plan and predict the outcome of the disease.

The results of this study suggested that there was no significant linear correlation between lymphocyte subsets and cytokines. By analyzing the differences of lymphocyte subsets and cytokines in peripheral blood between the mild and severe patients, we found that only CD4 + T, CD8 + T, IL-6, IL-10 had statistical significance between the mild and severe groups, suggesting that the immunosuppression of severe patients with 2019-nCoV infection was more obvious, which was consistent with the opinions of many experts.^5, 6^ For several results of this study, for example, the proportion of patients with an IL-6 value of 0 in the mild group was as high as 55.88%, there was no significant difference in terms of IL-17, TNF, IFN, and IL-4 between the two groups of patients, both B cell and NK cell decreased in different degrees in two groups of patients, the possible reasons are as follows. (1)The conclusion of this study is a true reflection of the characteristics of 2019-nCoV itself; (2) In the early stage of 2019-nCoV infection, due to the strong variability and good concealment of the virus, it cannot be quickly recognized by the body; (3) 2019-nCoV releases certain special factors after entering the body and interferes with the body’s initiation of a specific immune response; (4) Other reasons that have not yet been confirmed.

This study has several limitations. Firstly, as the largest diagnosis and treatment center for patients with NCP in Chongqing area, our hospital has more than 123 patients so far. The sample size was relatively small compared with Wuhan, where the disease originated, which may have some impact on the statistical results. But on the whole, the number of patients in this area was in the middle level for other parts of the country except Wuhan, and the research results were relatively reliable. Secondly, the humoral immunity level of the included patients was not monitored, so there was a certain deficiency in the evaluation of the immune system. Thirdly, due to the large-scale outbreak of the epidemic restricting the flow of people, data on healthy patients are lacking as blank controls.

In future studies, data will be collected from healthy patients as blank controls to further explore the predictive value of peripheral blood lymphocyte subsets and cytokines for patients with 2019-nCoV infection. At the same time, we will cooperate with other designated treatment units to carry out multicenter research to include more confirmed 2019-nCoV infected patients, expand the sample size, and design more rigorous randomized controlled trials. In addition, we will strengthen the follow-up of patients who are cured and discharged, and regularly detect the patient’s peripheral blood lymphocyte subsets and cytokines.

## Data Availability

The datasets generated and analyzed during the current study are available from the corresponding author on reasonable request.

## Acknowledgement

We sincerely thank Lian Guo, director of the Teaching Department of the Chongqing Three Gorges Central Hospital, and Chunhui Lang, director of the Foreign Affairs Department of Scientific Research, for their great support to this subject.

## Author Contribution

J.L. had the idea for and designed the study and had full access to all data in the study and take responsibility for the integrity of the data and the accuracy of the data analysis. S.W., S.F., X.Z., Q.X., K.X., J.X., B.Z. and Y.C. contributed to writing of the report. L.G. and C.L. contributed to critical revision of the report. Q.Y., Z.Y. and M.Q. contributed to the statistical analysis. All authors contributed to data acquisition, data analysis, or data interpretation, and reviewed and approved the final version.

